# Genetically determined alterations in inflammation and infection-associated genes are associated with hydrocephalus in patients of African Ancestry

**DOI:** 10.1101/2025.06.05.25329021

**Authors:** Andrew T. Hale, Jing He, Lisa Bastarache, Albert M. Isaacs

## Abstract

**Background and Objectives:** The genetic mechanisms underlying hydrocephalus (HC) risk and pathogenesis are diverse and complex. While many human genetics studies of HC have been performed in largely European ancestry patients, the extent to which these genetic mechanisms are conserved in African ancestry populations is not known. Here we apply complementary and convergent human genetics and functional genomics approaches to identify genes and pathways implicated with HC risk in patients of African ancestry.

**Methods:** We perform a transcriptome-wide association study (TWAS) in pediatric patients of African ancestry with HC across 3,288 individuals (46 cases and 3,242 controls). Gene set enrichment analysis (GSEA) and functional genomics were performed to identify genetically determined pathways conferring HC risk and potential mechanisms of implicated genes, respectively.

**Results:** After multiple-testing correction (false discovery rate< 0.05) across all genes tested, we identify decreased expression of *TMEM208* meeting experiment-wide statistical significance. *TMEM208* met the highly stringent Bonferroni threshold for statistical-significance based on the total number of genes tested (OR= 2.07, p< 5.22x10^-8^) indicating that *TMEM208* is a transcriptome-wide predictor of HC. STRING analysis identified co-expression and protein-protein interaction networks associated with TMEM208 enriched for infection and inflammation-associated genes, providing the basis for mechanistic, hypothesis-driven experiments to delineate the role of this gene in conferring HC risk. Finally, GSEA of all nominally associated genes (p< 0.05) revealed a marked association with genes regulating inflammatory and infection-related processes.

**Conclusions:** We perform the first systematic genetic study of HC in patients of African ancestry. We identify genetically determined alterations in inflammation and infection-associated genes underlying hydrocephalus in patients of African ancestry. Co-evolution of humans and pathogen-imposed selection pressures have differentially shaped the genetic etiology of HC across ancestral populations.

## Introduction

Hydrocephalus (HC) is a failure of brain and cerebrospinal fluid (CSF) homeostasis often associated with dilation of the CSF-filled ventricles (ventriculomegaly) and leads to increased intracranial pressure. There are both genetic^1–3^ and acquired (i.e. post-infectious, post-hemorrhagic, etc.) etiologies of HC,^4^ where infection is the most common cause of HC globally,^5^ and disproportionately impacts non-European ancestry patients. Specifically, The incidence of HC in Sub-Saharan Africa (> 50% of the world’s HC cases) is largely due to infection-related cases.^6^ However, we lack of mechanistic understanding of HC, which has impeded progress towards pharmacological options.^7, 8^ One limitation to identifying and implementing pharmacologic therapies for HC is that systematic genetic studies of HC have only been performed in predominantly European ancestry cohorts.^1, 2, 9^

The co-occurrence of infection (systemic or neurological) and pleiotropic relationship with HC is well-documented.^10, 11^ This is perhaps not surprising since the population genetic architecture of African populations, where HC disease burden is highest and driven in part due to host-pathogen evolution, is enriched for genetic loci involved in infection.^12^ While recent efforts have been made to understand the influence of host genetic architecture on infectious disease (ID) susceptibility and related traits,^13^ the extent to which host genetic architecture underlying IDs is related to neurodevelopmental outcomes (i.e., HC), for which infection is a contributing factor in select cases, is lacking. In addition, there is emerging evidence that inflammatory pathways may play a broader role in conferring risk to HC even in the absence of a clear infectious etiology.^4^ Critical to understanding the relationship between HC and infection is first to understand the genetic underpinnings of HC.^14^ We believe human genetic studies of HC in peoples of African ancestry, where disease burden is highest and where host genomes have been differentially shaped by host-pathogen interactions and selection pressures, will enable elucidation of precise molecular mechanisms and identify putative therapeutic targets.

Transcriptome wide association studies (TWAS) which rely on identification of gene-based associations, rather than individual single nucleotide polymorphism (SNP) identification, have been recently applied to HC.^2^ TWAS can infer molecular mechanisms of the gene-based association owing to directionality measure of association with the phenotype of interest.^15^ Moreover, owing to reduced multiple-testing burden, TWAS benefits from a lower p-value threshold (thousands of genes vs. millions of SNPs) and typically relies on aggregation of common variants, compared to whole-exome sequencing or exome-array chips which are best at identifying *de novo* or rare variations (Figure 1A). We recently conducted a TWAS of HC and identified maelstrom (*MAEL*) in the cortex as the gene-tissue pair reaching experiment-wide significant association.^2^ This finding was supported by complementary and convergent functional genomics approaches, but was limited to patients of European ancestry. Here we apply similar methodology and perform a TWAS of patients with HC of African ancestry. Owing to limited availability of population genetic atlases in many African populations,^16^ TWAS is an attractive approach since this method relies on aggregation of SNPs across a gene and its regulatory region. Here, to our knowledge, we perform the first systematic genetic study of HC in patients of African ancestry. We identify *TMEM208* as a transcriptome-wide predictor of HC in African ancestry patients and implicate genetic variations in infection and inflammatory-related genes underscoring HC, even in the absence of infection as a preceding factor. Our data highlight the role of genetically encoded risk factors in infection and inflammation-related genes contributing to HC in patients of African ancestry.

**Figure 1.**
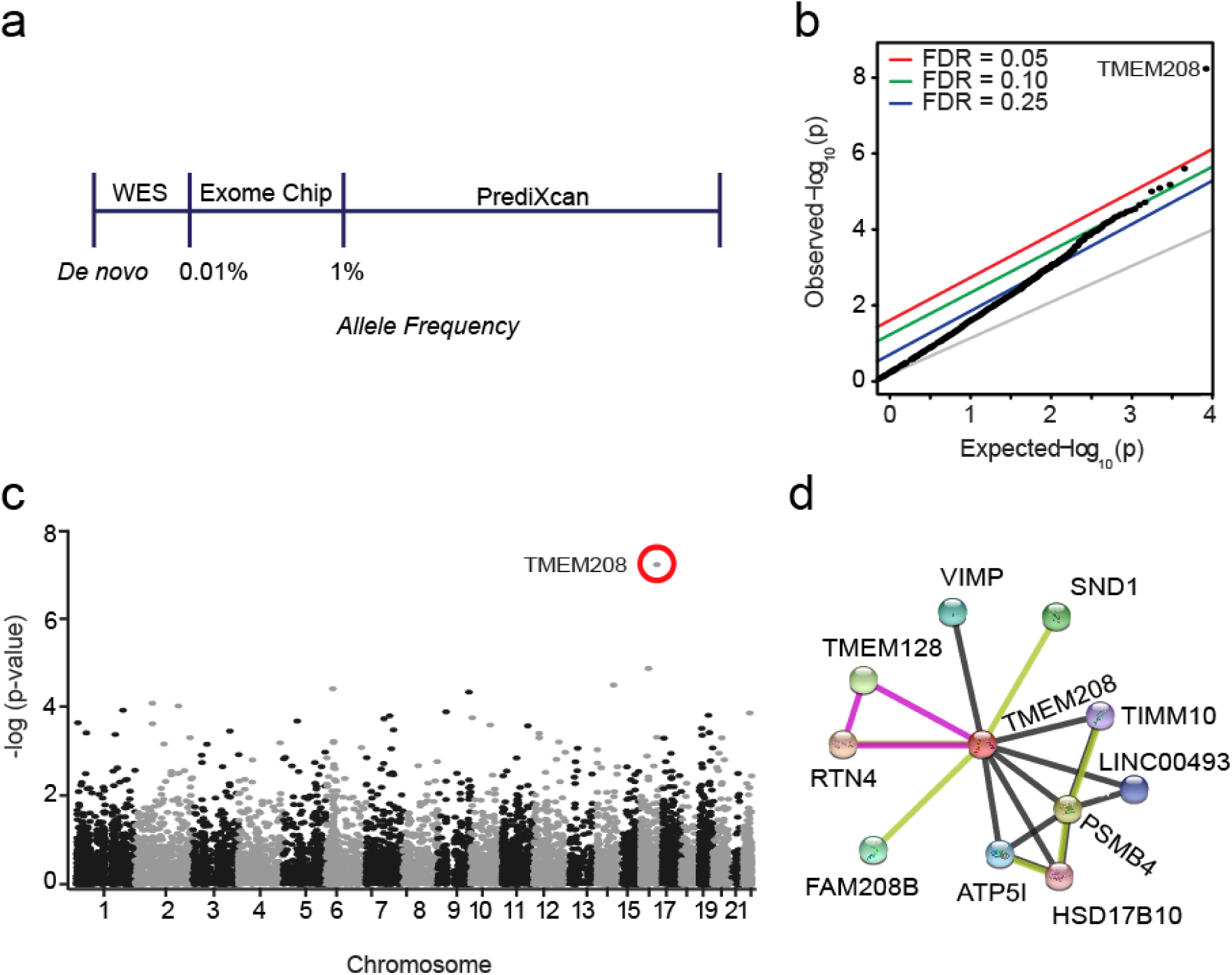
Transcriptome-wide association study (TWAS) of pediatric hydrocephalus in patients of African ancestry. (A) Overview of approaches to understand the genetic basis of disease, where whole-exome sequencing (WES), exome chip arrays, and transcriptome-wide association studies (TWAS, using PrediXcan) are aimed at understanding rare, common, and gene-regulatory variants, respectively. (**B**) Quantile-Quantile (Q-Q) plot demonstrating a significant association between *TMEM208* and hydrocephalus after correction using Benjamini-Hochberg false-discovery rate (FDR). *TMEM208* remains significant (adjusted p<0.05) after Bonferroni adjustment for the number of gene-tissue pairs tested in the study. **(C)** Manhattan plot demonstrating the chromosomal location of TMEM208 compared to other genes across the genome. **(D)** STRING analysis revealing TMEM208 interaction network derived from various lines of evidence: experimental determined (pink), text mining (light green), and co-expression (black). The individual proteins shown in the diagram are predicted functional partner.

## METHODS

### Patient Cohort

BioVU (DNA biobank linked to an EHR database) is comprised of the synthetic derivative (SD), a deidentified EHR which enable comorbidity estimates detailed below. For genetic analysis, BioVU contains whole-genome genetic information for 31,987 individuals which we apply here. Detailed information on the construction, utilization, ethics and policies of the BioVU resource is described elsewhere.^17^ The BioVU samples were included in the downstream joint-tissue PrediXcan analyses of the ID phenotypes. Genomic ancestry was quantified using principal components analysis as previously described.^18, 19^ We identified patients with a diagnosis of HC (Phecode: 331.1, phewascatalog.org^20, 21^) who have undergone permanent CSF diversion (VP shunt, ETV, or ETV/CPC).

### Transcriptome wide association study

Our TWAS included patients with HC of African ancestry (n= 46) and non-disease controls (n= 3,242). These analyses were performed as previously described.^2^ Briefly, we utilized PrediXcan,^22^ a TWAS method that estimates the genetic component of gene expression using imputation models derived from GTEx.^23–25^ This approach relies on logistic regression using the genetically-determined component of gene expression as independent variable and disease status where sex and age are covariates.

### Gene Set Enrichment Analysis

We hypothesized that genes identified through TWAS, which relies exclusively on the genetic component of gene expression and avoids potential environmental and technical confounding components of gene express analysis, would enable identification of pathways and biological processes underscoring genetic risk of HC. We used as input the genes identified through TWAS (p<0.05) with HC. Gene Set Enrichment Analysis (GSEA) ^26, 27^ was performed by determining enrichment of input genes in the curated gene sets in the Molecular Signatures Database (Broad Institute). Enrichment was determined using Benjamini-Hochberg adjusted p<0.05 in each tissue. Experiment-wide significant gene sets were identified using Bonferroni adjustment (adjusted p<0.05). STRING analysis was performed as previously described.^28^

## RESULTS

African genomes are among the most complex and diverse in the world, with an estimated ∼2,200 unique genetic diaspora.^29^ Owing to the burden of infectious diseases on the African continent and complex host-pathogen interactions over evolution, genetic loci involved response to infectious pathogens are likely to be under natural selection. Such signatures of selection may result from local, regional, and continental adaptations and determine the gene-level associations with human disease. Here we perform a TWAS in pediatric patients of African ancestry with HC across 3,288 individuals (46 cases and 3,242 controls). This is an admixed African American cohort of predominantly West African decent, which presents unique statistical and methodological challenges, as the combined effect of admixture and environment may lead to spurious associations and loss of statistical power. While our sample size for African-ancestry individuals with HC is small, which undoubtedly affects discovery power, our choice of methodology (TWAS by PrediXcan) enables a relaxed burden for multiple-testing correction owing to a lower number of statistical tests performed.

Despite these limitations, we identified transmembrane protein (*TMEM208*) as meeting the highly stringent Bonferroni threshold for statistical significance (*B_onferroni_* < 0.05) and false discovery rate (FDR) with HC in the African-ancestry individuals (Figure 1B-C). *TMEM208* has been shown to be a critical regulator of autophagy and endoplasmic reticulum (ER) stress,^30^ both critical mechanisms underlying cellular response to infection and injury. In addition, *TMEM208* has been shown to regulate cell polarity in *Drosophila melanogaster*, a common implicated cellular mechanism underlying HC.^31^ However, the role of *TMEM208* in mammalian systems is largely uncharacterized. Notably, this *TMEM208* shows highly African population-specific associations (p<0.001) with HC-related phenotypes in BioVU, including cerebral edema, cerebral degeneration, and other immunological findings, all meeting multiple-testing correction thresholds (Table 1).

**Table 1.**
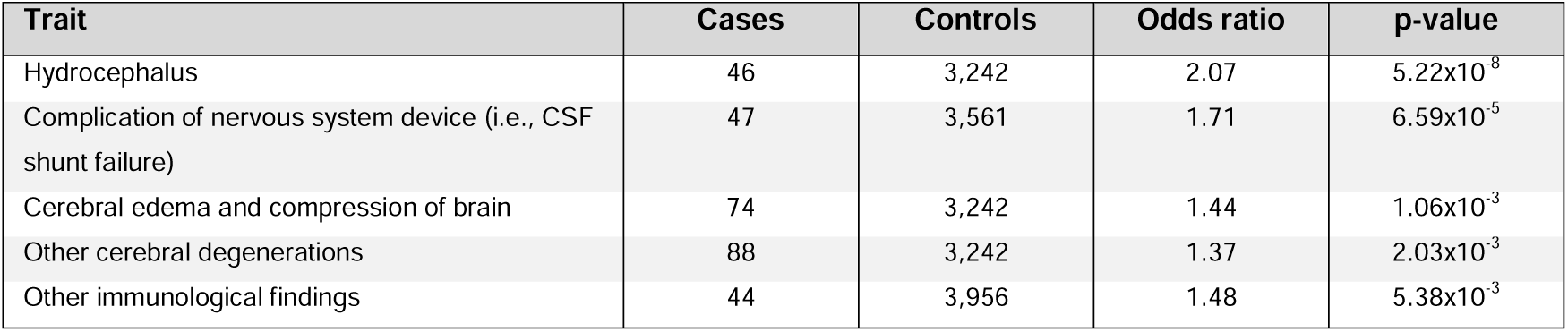
Phenotypes associated with genetically determined component of TMEM208 gene expression.

To further understand potential mechanisms by which TMEM208 may lead to HC, we performed STRING analysis,^32^ which estimates TMEM208 protein interactors based on experimentally determined, text mining, and co-expression levels of evidence. This approach identified 10 proteins in the TMEM208 interactome (Figure 1D). Using these 10 genes as input for gene-set enrichment analysis (GSEA), we identified a significant enrichment of immune-related genes in TMEM208-interacting proteins (Table 2). These data suggest that underlying alterations in infection-related gene networks can underlie susceptibility for hydrocephalus and CSF shunt failure.

**Table 2.** Functional enrichment in network of TMEM208-associated pathway identified by STRING analysis.

| GSEA Gene Set Description | False Discovery Rate (FDR) |
| --- | --- |
| **Biological process (Gene Ontology)** |  |
| Negative regulation of inflammatory response to antigenic stimulus | 0.015 |
| Regulation of inflammatory response to antigenic stimulus | 0.023 |
| Inner mitochondrial membrane organization | 0.048 |
| **Cellular component** |  |
| Mitochondrial protein complex | 0.048 |
| **KEGG Pathways** |  |
| Alzheimer's disease | 0.035 |

Intriguingly, genetically determined expression of *TMEM208* was also the most significant gene-level association with (‘failure of a CNS device,’ i.e., CSF shunt failure, Table 1), suggesting *TMEM208* may confer both disease and treatment failure risk for hydrocephalus. Since immune cell recruitment, accumulation, and function are factors associated with CSF shunt failure,^33^ this finding is consistent with our STRING analysis of the TMEM208 network. Indeed, all such neurological trait associations with *TMEM208* were detected in African-ancestry individuals (Table 1) and none in European-ancestry individuals, despite the larger sample size (and thus higher statistical power) for the latter. Moreover, *TMEM208* was not associated with HC in European patients.^2^

In an effort to understand the mechanisms contributing to HC implicated risk genes in patients of African ancestry patients, we performed enrichment analysis using the using the list of nominally associated HC genes (n=152 genes p< 0.05) as input). We identified immune-related genes (Table 3, Benjamini-Hochberg adjusted p=0.019), as curated in the Genetic Association Database,^34, 35^ as the leading gene set, suggesting the presence of bonafide signals in the input genes. Moreover, GSEA demonstrated significant enrichment for a number of immunologic gene sets (FDR<10^-6^, Table 4) among the HC-associated genes in patients of African ancestry. These data identify infection and inflammatory-associated genes underlying HC, even in the absence of infection as the inciting factor for the disease.

**Table 3.** Infection-related genes associated with hydrocephalus in African ancestry individuals.

| Gene |
| --- |
| ABCA7, ABCF1, ABHD16A, ACOXL, ACP5, ADAMTS10, AGER, ANAPC4, ANXA11, ATF6B, BAG6, BCL6, BICD1, BID, C4B, C6orf48, C7, CASP3, CD2, CD69, CDC42BPA, CFH, CHML, CHUK, CLN3, CNIH3, COL18A1, COMMD10, COX7C, CPAMD8, CPLX1, CREB1, CSF1R, CTNNA2, DCN, DDX20, DGKH, DHX16, DKK1, DLD, EFCAB11, ENDOU, EPS8L2, F2, F5, FAM13C, FASLG, FCAR, FCER2, FGFR4, FTO, FYN, GABRB2, GALNTL6, GNA12, GNRH1, GPANK1, HECA, HECW1, HERPUD2, HIF1A, HIGD1C, ICAM1, IL12A, IL18RAP, IL20RB, IL36G, INPP4A, INSR, ITGA4, ITGAV, ITGAX, ITGB7, KATNB1, KIR2DL1, KLRC2, LCE3A, LCE5A, LRRTM1, MACROD2, MALT1, MAPK14, MAPK3, MDM2, MRC1, MSANTD2, MSH5, MTRR, MTTP, NLRP1, NR2C1, NRDE2, OSMR, PALD1, PON2, PPM1L, PPP2R2C, PRKCSH, PRRC1, PRRC2A, PRSS16, PSMB1, PSMG1, PTCH1, PTPN6, PVR, RAB3C, RAC2, RAD54B, RNASE2, RNASE3, RNF5, ROM1, RUNX3, S100B, SELL, SFTA2, SH3TC2, SHBG, SLC22A6, SLC25A27, SLC39A12, SMARCE1, SMUG1, SPN, STAT3, SULF2, SV2B, SYT14, TFR2, TH, THBS4, TMEM135, TMEM18, TMTC2, TNFAIP6, TNFRSF25, TPPP, TRIM39, TUBAL3, UNC13B, UTS2, VARS2, WDFY4, WISP1, WNT3, WRN, XRCC5, ZFHX3, ZNF383, ZNF385B, ZSCAN31 |

**Table 4.** Gene Set Enrichment Analysis (GSEA) on nominally significant (p< 0.05) gene-level associations with hydrocephalus in individuals of African ancestry.

| GSEA Gene Set Description | P value | False Discovery Rate (FDR) |
| --- | --- | --- |
| **Hallmark gene sets** |  |  |
| Genes defining inflammatory response | 7.54x10<sup>-6</sup> | 3.77x10<sup>-4</sup> |
| Genes encoding components of blood coagulation system; also upregulated in platelets | 2.13x10<sup>-5</sup> | 5.31x10<sup>-4</sup> |
| Genes defining late response to estrogen | 1.46x10<sup>-4</sup> | 1.22x10<sup>-3</sup> |
| Genes up regulated in response to interferon gamma | 1.46x10<sup>-4</sup> | 1.22x10<sup>-3</sup> |
| Genes important for mitotic spindle assembly | 1.46x10<sup>-4</sup> | 1.22x10<sup>-3</sup> |
| Genes up regulated through activation of mTORC1 complex | 1.46x10<sup>-4</sup> | 1.22x10<sup>-3</sup> |
| Genes up regulated by activation of hedgehog signaling | 3.08x10<sup>-4</sup> | 2.19x10<sup>-3</sup> |
| Genes mediated programmed cell death (apoptosis) by activation of caspases | 3.89x10<sup>-4</sup> | 2.19x10<sup>-3</sup> |
| Genes upregulated during transplant rejection | 5.70x10<sup>-4</sup> | 2.19x10<sup>-3</sup> |
| Genes encoding components of the complement system, a component of the innate immune system | 5.70x10<sup>-4</sup> | 2.19x10<sup>-3</sup> |
| **Chromosomal position gene sets** |  |  |
| Genes in cytogenetic band chr4q34 | 2.16x10<sup>-6</sup> | 7.04x10<sup>-4</sup> |
| Genes in cytogenetic band chr19p13 | 1.46x10<sup>-5</sup> | 1.76x10<sup>-3</sup> |
| Genes in cytogenetic band chr8q24 | 1.62x10<sup>-5</sup> | 1.76x10<sup>-3</sup> |
| Genes in cytogenetic band chr6p21 | 3.74x10<sup>-4</sup> | 3.04x10<sup>-2</sup> |
| **Cis-regulatory motif gene sets** |  |  |
| Genes containing the highly conserved motif M13 CTTTGT | 1.47x10<sup>-22</sup> | 1.23x10<sup>-19</sup> |
| Genes containing the highly conserved motif M6 GGGCGGR | 5.65x10<sup>-16</sup> | 2.36x10<sup>-13</sup> |
| Genes containing the highly conserved motif M17 AACTTT | 1.27x10<sup>-15</sup> | 3.54x10<sup>-13</sup> |
| Genes containing the highly conserved motif M24 GGGAGGR | 2.24x10<sup>-13</sup> | 4.69x10<sup>-11</sup> |
| Genes containing the highly conserved motif M12 CAGGTG | 7.37x10<sup>-13</sup> | 1.23x10<sup>-10</sup> |
| Genes containing the highly conserved motif M3 SCGGAAGY | 6.71x10<sup>-12</sup> | 7.09x10<sup>-10</sup> |
| Genes containing the highly conserved motif M15 CAGCTG | 6.76x10<sup>-12</sup> | 7.09x10<sup>-10</sup> |
| Genes containing the highly conserved motif M60 TTGTTT | 6.78x10<sup>-12</sup> | 7.09x10<sup>-10</sup> |
| Genes containing the highly conserved motif M56 GGGTGGRR | 1.42x10<sup>-11</sup> | 1.32x10<sup>-9</sup> |
| Genes containing the highly conserved motif M5 GATTGGY | 1.83x10<sup>-11</sup> | 1.53x10<sup>-9</sup> |
| **Gene Ontology gene sets** |  |  |
| GO_ESTABLISHMENT_OF_LOCALIZATION_CELL | 8.45x10<sup>-25</sup> | 5.00x10<sup>-21</sup> |
| GO_REGULATION_OF_TRANSPORT | 3.06x10<sup>-24</sup> | 9.05x10<sup>-21</sup> |
| GO_PROTEIN_LOCALIZATION | 1.18x10<sup>-23</sup> | 2.33x10<sup>-20</sup> |
| GO_RIBONUCLEOTIDE_BINDING | 2.77x10<sup>-23</sup> | 4.10x10<sup>-20</sup> |
| GO_ENZYME_BINDING | 3.99x10<sup>-23</sup> | 4.72x10<sup>-20</sup> |
| GO_INTRACELLULAR_VESICLE | 2.09x10<sup>-21</sup> | 2.06x10<sup>-18</sup> |
| GO_PHOSPHATE_CONTAINING_COMPOUND_METABOLIC_PROCESS | 6.95x10<sup>-21</sup> | 5.88x10<sup>-18</sup> |
| GO_ADENYL_NUCLEOTIDE_BINDING | 9.28x10<sup>-21</sup> | 6.87x10<sup>-18</sup> |
| GO_HOMEOSTATIC_PROCESS | 1.70x10<sup>-20</sup> | 1.12x10<sup>-17</sup> |
| GO_ESTABLISHMENT_OF_PROTEIN_LOCALIZATION | 4.39x10<sup>-20</sup> | 2.60x10<sup>-17</sup> |
| **Immunologic gene sets** |  |  |
| Genes up regulated in activated CD4 T cells | 2.01x10<sup>-10</sup> | 4.89x10<sup>-7</sup> |
| Genes upregulated in activated CD4 T cells by anti-CD3 and anti-CD28 then stimulation with IL-12 | 2.01x10<sup>-10</sup> | 4.89x10<sup>-7</sup> |
| Genes upregulated in macrophages exposed to B. Malayi vs. macrophages exposed to M. tuberculosis | 1.35x10<sup>-9</sup> | 2.20x10<sup>-6</sup> |
| Genes up-regulated in bone marrow-derived dendritic cells stimulation with CSF2 and 1,3 beta-D-oligoglucan | 7.34x10<sup>-9</sup> | 3.49x10<sup>-6</sup> |
| Genes down regulated in STAT3 knockout CD4 T cells treated with TGF-β | 7.95x10<sup>-9</sup> | 3.49x10<sup>-6</sup> |
| Genes up regulated in dendritic cells stimulation with TLR7 agonist | 8.60x10<sup>-9</sup> | 3.49x10<sup>-6</sup> |
| Genes up regulated in NFATC1 knockout B lymphocytes stimulated with anti-IgM | 8.60x10<sup>-9</sup> | 3.49x10<sup>-6</sup> |
| Genes up regulated in CD8 T cells over-expressing ID3 | 8.60x10<sup>-9</sup> | 3.49x10<sup>-6</sup> |
| Genes up regulated in LSK vs. CD8 T cells | 8.60x10<sup>-9</sup> | 3.49x10<sup>-6</sup> |
| Genes downregulated in BCL6 heterozygous knockout marginal zone B lymphocytes | 8.60x10<sup>-9</sup> | 3.49x10<sup>-6</sup> |

## DISCUSSION

We present the first systematic human genetic study of HC in patients of African ancestry. Our data identify *TMEM208* as a transcriptome-wide predictor of HC and CSF shunt failure in patients of African ancestry, despite a modest sample size. These data are corroborated by unbiased functional genomics analysis implicating genes involved in infection and inflammation as potentially causative. Owing to the genetic selection pressures faced by African populations over time as well as co-evolution with pathogens and other selection pressures, our data suggest that genetic risk for HC has been differentially shaped across European and African ancestral populations. These data serve as a foundation for larger genetic studies of HC in non-European populations and for future mechanistic studies aimed at understanding the role of infection/inflammation in germline HC genetic risk.

The co-occurrence of infection (systemic or neurological) and HC in both the developed and developing world is well-documented.^10, 11^ While post-infectious HC has largely been considered a disease without a germline genetic component, our data suggest that African populations may have genetically encoded risk to HC in infection/inflammatory pathways, even in cases where infection is not the cause of HC. Data from murine models suggest that pathways triggered by pathogen exposure may also contribute to post-hemorrhagic HC.^4, 36^ In addition, many cases of infection-related HC occur in the absence of “active” bacterial infection in the CSF (and where complete viral characterization are not always assessed) and in the presence of hemorrhage (an independent risk-factor for hydrocephalus).^37, 38^ Moreover, precise identification of infectious organisms remains a challenge, but advances in molecular engineering and reverse genomics have provided strategies to identify and culture these pathogens,^39, 40^ which may increase our understanding of host-pathogen interactions and diseases.^41, 42^ For example, proteogenomic studies of post-infectious HC in Ugandan infants revealed a number of pro-inflammatory pathways associated with the disease.^39^ This is perhaps not surprising since the population genetic architecture of African populations, driven in part due to co-evolution of pathogens, is enriched for genetic loci involved in infection.^12^ Furthermore, pan-microbial analysis of CSF and blood identified a novel, previously thought not to cause human disease, strain of *Paenibacilus thiaminolyticus* (as well as cytomegalovirus) as key contributors post-infectious HC.^43^ It is certainly possible that certain germline genetic variations confer shared risk to both HC and infection risk in this founder population(i.e., genetic theory of infectious disease),^44^ but not among other populations.

The most common treatment of HC, insertion of a CSF shunt, is often complicated by infection,^45^ but has many contributing factors.^46^ Recent evidence using 16S rRNA sequencing of CSF from patients with shunt infection demonstrates a highly polymicrobial profile among individuals with shunt infection, but the precise causative pathogen is not always known as broad spectrum antibiotics are deployed and narrowed upon subsequent culture information.^47^ Furthermore, the extent to which host genetic variation influences inflammatory response at the ependymal surface adjacent to the shunt vs. susceptibility to IDs that lead to CSF shunt failure are not known. Since HC and CSF shunt failure risk is multi-factorial,^37, 48–50^ a comprehensive study of phenotypic and genetic factors across the ID phenome, HC development, and treatment failure (e.g., CSF shunt failure, scarring of the ETV ostomy site,^51^ or even emerging contributions of the choroid plexus (ChP) to mediating infection and inflammation^52^ coupled with the unknown biological and mechanistic basis of ChP cauterization). The identification of a single gene conferring both risk to HC disease, and independently, to CSF shunt malfunction suggest that germline genetic factors should be taken into consideration for these patients.

Our study also highlights the importance of linking electronic health records (EHR) information to genetic data. Much of the public and global health burden of IDs is in resource-poor settings and disproportionately affects children,^53, 54^ underscoring the range of environmental risk factors (hygiene, age, and socioeconomic status, among others) that may be contributing to ID risk. Because of the disproportionate burden of IDs in resource-limited settings where health records may not be easily obtained and screening/laboratory procedures differ, a systematic genetic study of predisposition to ID and HC remains logistically challenging. Defining infection (i.e., presence of growth in culture, symptomatology of the patient, systemic inflammatory response, etc.) is also complicated and highly region/setting specific. While most genetic studies collate large numbers of samples for individuals traits, this approach may not be possible in these contexts, necessitating evolving strategies.

We present a systematic human genetic study of HC in patients of mixed African ancestry and identify TMEM208 reaching experiment-wide significance. While our sample size is modest and require validation in an independent cohort, these data highlight the utility of TWAS in identifying the genetic basis of rare disorders where single genetic variants with large effect are unlikely to explain the genetic contributions. Availability of African ancestry genomic data remains a challenge, requiring prospective collection and genetic analysis of diverse peoples. Because of the considerable degree of genetic variations, sequencing approaches, rather than chip-based arrays, will yield the most important insights. Nonetheless, our data provide proof of principle that population ancestry differentially impacts genetic risk of HC and prompts consideration for large-scale human genetics studies of HC in non-European populations.

## Data Availability

All data produced in the present work are contained in the manuscript.

